# Navigating housing independence: transitions out of the parental home of young Australians with and without disability

**DOI:** 10.64898/2026.02.11.26346107

**Authors:** Tess Bright, Glenda Bishop, Kate Mason, Alex Sully, David Gurrin, Helen Dickinson, Anne Kavanagh, Zoe Aitken

## Abstract

Young people are increasingly remaining in the parental home for longer - a trend associated with poorer mental health. There is little evidence on this transition for young people with disability. We used three waves of the Australian Census Longitudinal Dataset, a 5% sample of linked Census records. Two analyses compared transitions between 2011-2016 and 2016-2021 among people 15-34y living with parents at baseline with complete data on disability and housing. The proportion of people no longer living with parents at follow-up was calculated, comparing people with and without disability, along with absolute and relative inequalities. Young people with disability were half as likely to leave the parental home as their peers without disability. Inequalities were greatest for people 25-29y (relative difference 0.41 (95%CI 0.36-0.45), living outside major cities (0.48, 0.44-0.52), or with higher income (0.53 (0.47-0.59). Patterns were consistent over time. Targeted supports are needed to enable independent living.

**Points of interest:**

- We found that less people with disability leave the parental home than people without disability
- We also found the gap between people with and without disability was biggest outside major cities.
- This may mean people with disability in rural, regional and remote areas find it more difficult to move out of home
- Better housing and income supports are needed to help young people with disability live in the way they choose

## Introduction

Article 19 of the United Nations Convention on the Rights of People with Disability (UNCRPD) states that people with disability have the right to choose a safe, secure and community-based home on an equal basis with others (1). Despite Australia being a signatory to the UNCPRD it has not achieved this vision with current evidence demonstrating that people with disability experience significant housing disadvantage. Compared to people without disability, people with disability in Australia are much more likely to live in unaffordable (2, 3) or poor quality housing (4), and are at increased risk of homelessness (5, 6). People with disability also experience disadvantage across a range of other socio-economic areas including lower rates of employment and workforce participation, lower income, and higher costs of healthcare, which further compounds housing disadvantage. These inequalities reinforce a cycle of disadvantage in housing and further limit housing choice. Since housing is a key social determinant of health this inequity is a critical issue.

There are more than 300,000 young people (15-34 years) with disability in Australia (7). It is important to recognise that this is a diverse group – with disparate needs in terms of housing and support. This age group represents a time of transition to independence, as young people engage in post-school education, employment, and move away from the parental home. Moving away from the parental home is not only an important marker of independence, but there are also potential health benefits. An Australian study by Howard et al. (2023), found that young people who co-reside with parents had poorer mental health than those who lived independently (8). However, experiences of living with parents vary, and cultural expectations shape when and how young people leave home. Some young people benefit from strategic co-residence (e.g. avoiding rent while saving for a home deposit). Yet for many young people, delaying transition from the parental home is the only feasible option given the scarcity of affordable housing options. Therefore the mental health impacts may vary between different groups.

A large body of evidence has established that housing quality, insecurity, and affordability are closely linked to health outcomes for people of all ages (9–13). In the context of the current global housing affordability crisis, housing affordability is receiving more attention as a determinant of health, including mental health among young people, with several recent systematic reviews and longitudinal studies finding strong evidence of this (14–16). Conversely, access to safe, stable, and affordable housing is associated with a range of positive health outcomes, including reduced disease burden from cardiovascular disease, asthma or injury, and increased quality of life (17). Housing is also a key factor in the International Classification of Functioning Disability and Health (ICF) framework, which acknowledges that where a person lives influences their ability to participate and function in society (18). Safe, stable, and affordable housing can therefore act as a foundation for participation in education and employment and improved social inclusion.

Young people in high-income countries are increasingly remaining living with their parents well into early adulthood (19). In Australia, with approximately half of males (54%) and females (47%) aged 18-29 still living with their parents (20). Analysis of the 2020 Household, Income and Labour Dynamics in Australia (HILDA) survey found that the average age to move out of home is 24 years for males and 23 years for females, compared to 23 and 22 years, respectively, in 2002 (21). This delayed transition is linked to the broader housing affordability crisis, but also other factors, such as changing employment opportunities, education participation, and later life milestones such as marriage and having children. The transition out of the family home is not only later, but also more complex, with many young people returning to the parental home at various stages (22). However, a 2019 longitudinal analysis of HILDA data found that the higher proportion of young people living with their parents in recent years is driven by a lower proportion moving out rather than an increased proportion moving back into the parental home (23). Whilst there are published trends documented in the general population, less is known about how structural pressures intersect with disability. The challenges of leaving the family home are compounded for young people with disability due to socioeconomic disadvantage and the added complexity of some young people with disability requiring extra support or accessible homes. Evidence shows that people with disability want to live more independently but are unable to due to limited supply of affordable, accessible, and supported housing (24). Young people aspire to move out, with one survey finding 60% of emerging adults stating independent ownership was their ideal tenure type and 5% preferred living with parents/guardians (25). However, young people are often delayed by cost, a barrier which will disproportionately affect young people with disability (25, 26).

Challenges remain despite major reform in disability policy through the National Disability Insurance Scheme (NDIS). The NDIS aims to provide people with significant and permanent disability greater choice and control over their lives, including their living arrangements, by providing participants with individual budgets to purchase services they need. The NDIS funds a range of supports under the “home and living” category, including capital expenditure for home modifications or Specialist Disability Accommodation, as well as core and capacity building supports such as assistance to live independently (e.g. Supported Independent Living) or support to find and maintain a place to live. Although an important reform, most young people with disability are not eligible for home and living supports through the NDIS. Of the 5.5 million Australians with disability, 14% are NDIS participants, and only 7% of these receive intensive living support (7, 27, 28). A quarter of NDIS participants are aged 15-34 years – thus over 70% of young people with disability do not receive NDIS support (29, 30).

While existing literature has examined the housing challenges for people with disability, there is a lack of evidence on transitions from the parental home for young people with and without disability in Australia, and particularly whether there are disability-related inequities in these housing transitions. This is important in the context of the NDIS, and considering Australia being a signatory of the UNCRPD, both of which highlight housing choice as paramount for human rights and a central focus of disability policy reform in Australia. We do not know what housing transitions for young people with disability look like since the introduction of the NDIS and other policy reforms that have occurred at the same time as an escalation in housing costs. This article aims to fill this gap. This study aims to describe housing transitions out of the parental home for young people (15-34 years) with and without disability using data from the Australian Census Longitudinal Dataset (2011–2021), comparing two time periods (2011-2016 and 2016-2021) to assess changes over time. We then examined whether there were disability-related inequities, and whether these were associated with various demographic or socioeconomic characteristics.

### Research process

#### Datasets

This study uses data from Australian Census Longitudinal Dataset (ACLD) (31). ACLD uses a sample of the Census of Population and Housing to create an enduring longitudinal dataset through linking of records from successive Censuses. Four waves of Census data have contributed to the dataset: 2006 (wave 1), 2011 (wave 2), 2016 (wave 3) and 2021 (wave 4). A multi-panel framework is applied to maintain the ACLD sample. There are four ACLD panels, representing a sample of 5% of people enumerated in the Australian Census on the 2006, 2011, 2016, 2021 Census nights. ACLD commenced with a 5% panel sample from the 2006 Census, which was linked to subsequent Censuses using deterministic and probabilistic linkage techniques. Panel samples from the later Censuses were created using the same sampling techniques in order to maintain a linked sample of 5% of the population and to introduce new records to account for births, migration, and missing links in previous panels.

This study uses a longitudinal design, analysing linked cross-sectional data from three of the four ACLD waves - 2011, 2016, and 2021. We chose these years to examine trends prior to and after the introduction of the NDIS. Participants were selected from the ACLD based on their inclusion in the 2011, 2016, and 2021 Census datasets, with a sample drawn from individuals aged 15-34 years living in private dwellings in Australia. Individuals with missing data on key housing or disability variables were excluded. The sample was defined as people aged 15 to 34 years in the baseline wave (either 2011 or 2021) who: i) participated in two consecutive Censuses (i.e. 2011 and 2016, or 2016 and 2021); ii) have linked records; iii) were living with parents at “baseline” (i.e. 2011 and 2016); and iv) had complete data on disability, sociodemographic, and housing variables.

We undertook two sets of descriptive analyses to explore whether there have been changes in housing transitions for young people with and without disability over time, comparing transitions between 2011-2016 and 2016-2021.

Figure 1 show the sample selection as flowcharts; the definitions of the measures are provided below. The total sample included 80,012 people for the 2011-2016 analysis and 103,105 people for the 2016-2021 analysis.

**Figure 1:**
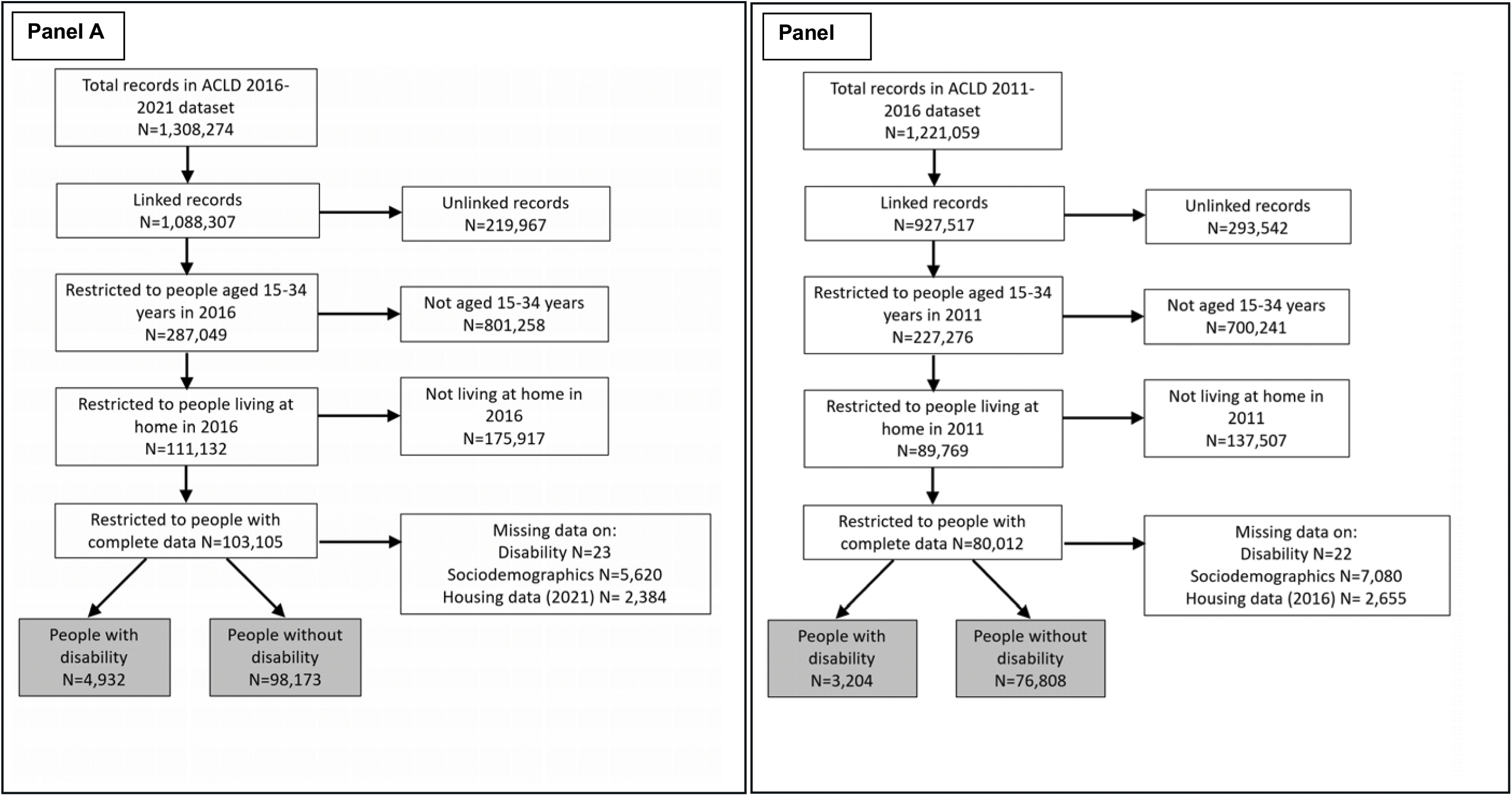
Flowchart showing sample selection for 2011-2016 (panel A) and 2016-2021 analyses (panel B).

#### Disability measure

Each Census includes four survey questions that the disability measure is derived from:

- Does the person ever need someone to help with, or be with them for self-care activities?
- Does the person ever need someone to help with, or be with them for, body movement activities?
- Does the person ever need someone to help with, or be with them for, communication activities?
- What are the reasons for the need for assistance or supervision?

Participants are defined as having a profound or severe core activity limitation if they have a need for assistance in their day-to-day lives in one or more of the three core activity areas of self-care, mobility, and communication because of: a long-term health condition (lasting six months or more), a disability (lasting six months or more), or old age. The Census does not include questions on specific disability types. For this study, a person was defined as having a disability if they met the above definition in either two waves (2011 or 2016; 2016 or 2021).

#### Parental co-residence status

A binary measure of ‘parental co-residence’ was derived from the Census “relationship in the household” variable – ‘living with parents’ or ‘no longer living with parents’. Relationship in the household (including grandchildren) describes the relationship of each person in a family to the family reference person. Where a person is not part of a family, their relationship to the reference person is described. Those classified as dependent student, or non-dependent child were coded as ‘living with parent(s)’. In addition, dependent student grandchildren, and non-dependent grandchildren were also classified as ‘living with parent(s)’. All others were classified as ‘no longer living with parent(s)’ provided they had complete data on housing tenure type (renter, owner).

#### Demographic and socio-economic measures

Demographic and sociodemographic variables at baseline (wave 1) were included in the analysis. Demographic variables included age, sex, Aboriginal and Torres Strait Islander status, country of birth of parents (Australia, major English-speaking country, other). Socio-economic characteristics were measured at the individual and area level. Individual variables included engagement in study or employment (derived variable based on engagement in full or part time study or employment), and personal income (derived and dichotomised into lowest 40% and highest 60%). Area level socio-economic characteristics were location (major city, outside major city), and Socio Economic Index for Areas (SEIFA) Index of Relative Socio economic Advantage and Disadvantage. This SEIFA index is constructed by the Australian Bureau of Statistics (ABS) using Census data and is defined “as people’s access to material and social resources, and their ability to participate in society” (32). The dimensions include income, education, employment, occupation, housing, and family structure. The index was dichotomised at the median to create a binary variable.

#### Statistical analysis

Descriptive analyses were conducted using Stata version 18.0 in August 2025 within the ABS DataLab to describe the Australian population of young people aged 15-34 years in terms of the demographic and socioeconomic variables outlined above. To compare housing transitions for people with and without disability, we calculated estimates of the proportion of people not living with their parents in the second wave, and 95% confidence intervals, stratified by disability status – restricted to people living with parent(s) at baseline (2011 or 2016). We also stratified by age, sex, engagement status, location, and personal income. We then calculated absolute and relative inequalities between people with and without disability, and 95% confidence intervals. Absolute inequality – a measure of inequality that focuses on the absolute difference between groups - was calculated by subtracting the proportion of people without disability not living with parents from the proportion of people with disability not living with parents. Relative inequality was calculated by dividing the proportion of people with disability living with parents by the proportion of people without disability living with parents. We did this for the overall estimates and for each of the sociodemographic variables mentioned above. We examined each inequality for overlapping 95% confidence intervals to assess differences between young people with and without disability.

An analysis of missing data was conducted by comparing proportions for those with and without missing socio-demographics and linked/unlinked records to determine the extent of possible selection bias.

#### Engagement with people with lived experience

As part of the study, we engaged with young people with disability to design the study and interpret the findings. The findings were discussed in collaborative sessions between the lead author and two lived experience research assistants. In these meetings, the emerging results were reviewed and discussed, and the research assistants’ reflections and lived expertise directly informed how the findings were interpreted and presented.

#### Ethical approval

Ethical approval was obtained from the University of Melbourne Human Research Ethics Committee (approval number: 30402). Data for this study was supplied by the ABS in the Person Level Integrated Dataset (PLIDA), 2021, Australian Census Longitudinal Dataset, accessed within the ABS Datalab, a secure online platform available to approved data users. All data presented in this paper was approved by the ABS prior to release from DataLab. We sought a waiver of consent for PLIDA data because the quantity of records (whole population data) makes this unfeasible, and the risk is low as the data is deidentified.

## Results

For the 2011-2016 cohort, at baseline in 2011 there were 3,204 people with disability representing 4.0% of the total cohort. For the 2016-2021 cohort, at baseline there were 4,932 people with disability (4.8% of cohort).

Table 1 describes the sociodemographic characteristics of people with and without disability at baseline. There were differences across all sociodemographic variables included in the analysis and patterns that were similar for both time periods. In 2016, the average age of people with disability was 21.6 years compared to 20.4 years for people without disability. A higher proportion of people with disability were male (61.4% [95% CI 60.7, 62.8] of people with disability vs. 53.3% [95% CI 53.0, 53.6] of people without disability); identified as Indigenous (5.9% [95% CI 5.3, 6.6] vs. 2.7% [95% CI 2.6, 2.8]); and were born in Australia (64.1% [95% CI 62.7, 65.4] vs. 52.8% [95% CI 52.5, 53.2]). A larger proportion of people with disability (62.0%; 95% CI 60.6, 63.4) were engaged in employment or education compared to people without disability (91.9%; 95% CI 91.7, 92.1). A higher proportion of people with disability had personal income in the lowest 40% of the distribution relative to people without disability (89.2% [95% CI 88.3, 90.0] vs. 70.8% [95% CI 70.5, 71.1]). A lower proportion of people with disability lived in a major city compared to their peers without disability (70.5% [95% CI 69.2, 71.8] vs. 79.0% [95% CI 78.7, 79.2]), and a higher proportion of people with disability had a SEIFA score in the lowest 50% (58.7% [95% CI 57.4, 60.1] vs 45.3 [95% CI 44.9, 45.6]).

**Table 1:** Population-weighted characteristics of young people aged 15-34 years who were living with parents in 2011 ACLD (n=80,012) and 2016 (103,105)

|  |  | 2011 |  | No disability |  | 2016 |  | No disability |  |
| --- | --- | --- | --- | --- | --- | --- | --- | --- | --- |
|  |  | Disability |  |  |  | Disability |  |  |  |
|  |  | N (%) | % (95% CI) | N (%) | % (95% CI) | N (%) | 95% CI | N (%) | 95% CI |
| **Mean age (mean, sd)** |  | 21.8 | 5.5 | 20.0 | 4.4 | 21.6 | 5.5 | 20.3 | 4.5 |
| **Age group** | 15-19 | 1390 (43.4) | (41.7, 45.1) | 42806 (55.7) | (55.4, 56.1) | 2206 (44.7) | (43.4, 46.1) | 50900 (51.9) | (51.5, 52.2) |
|  | 20-25 | 836 (26.1) | (24.6, 27.6) | 22216 (28.9) | (28.6, 27.6) | 1331 (27) | (25.8, 28.2) | 30306 (30.9) | (30.6, 31.2) |
|  | 26-29 | 574 (17.9) | (16.6, 19.3) | 8295 (10.8) | (10.6, 11.0) | 786 (15.9) | (14.9, 17.0) | 12036 (12.3) | (12.1, 12.5) |
|  | 30-34 | 404 (12.6) | (11.5, 13.8) | 3491 (4.6) | (4.4, 4.7) | 609 (12.4) | (11.5, 13.3) | 4931 (5.0) | (4.9, 5.2) |
| **Sex** | Male | 1983 (61.9) | (63.6, 64.2) | 41370 (53.9) | (53.5, 54.2) | 3030 (61.4) | (60.7, 62.8) | 52319 (53.3) | (53, 53.6) |
|  | Female | 1221 (38.1) | (36.4, 39.8) | 35438 (46.1) | (45.8, 46.5) | 1902 (38.6) | (37.2, 39.9) | 45854 (46.7) | (46.4, 47.0) |
| **Aboriginal and Torres Strait Islander** |  |  |  |  |  |  |  |  |  |
|  | Non-indigenous | 3063 (95.6) | (94.8, 96.3) | 74921 (97.5) | (97.4, 97.7) | 4642 (94.1) | (93.4, 94.7) | 95519 (97.3) | (97.2, 97.4) |
|  | Indigenous | 141 (4.4) | (3.7, 5.2) | 1887 (2.5) | (2.4, 2.6) | 290 (5.9) | (5.3, 6.6) | 2654 (2.7) | (2.6, 2.8) |
| **Country of birth of parents** |  |  |  |  |  |  |  |  |  |
|  | Australia | 2021 (63.1) | (61.4, 64.7) | 40679 (53) | (52.6, 53.3) | 3160 (64.1) | (62.7, 65.4) | 51875 (52.8) | (52.5, 53.2) |
|  | One or both English speaking | 503 (15.7) | (14.5, 17.0) | 11792 (15.4) | (15.1, 15.6) | 728 (14.8) | (13.8, 15.8) | 14630 (14.9) | (15.1, 15.0) |
|  | One or both non-English speaking | 680 (21.2) | (19.8, 22.7) | 24337 (31.7) | (31.4, 32.0) | 1044 (21.2) | (20.1, 22.3) | 31668 (32.3) | (31.7, 32.6) |
| **Engagement in study/work** |  |  |  |  |  |  |  |  |  |
|  | Not engaged | 1189 (37.1) | (35.5, 38.8) | 5660 (7.4) | (7.2, 7.6) | 1874 (38.0) | (36.7, 39.4) | 7955 (8.1) | (7.9, 8.3) |
|  | Engaged | 2015 (62.9) | (61.2, 64.6) | 71148 (92.6) | (92.4, 92.8) | 3058 (62.0) | (60.6, 63.4) | 90218 (91.9) | (91.7, 92.1) |
| **Individual income** | Highest 60% | 450 (14.0) | (12.9, 15.3) | 23809 (31.0) | (30.7, 31.3) | 533 (10.8) | (10, 11.7) | 28648 (29.2) | (28.9, 29.5) |
|  | Lowest 40% | 2754 (86.0) | (84.7, 87.1) | 52999 (69.0) | (68.7, 69.3) | 4399 (89.2) | (88.3, 90.0) | 69525 (70.8) | (70.5, 71.1) |
| **Location** | Major city | 2204 (68.8) | (67.2, 70.4) | 59547 (77.5) | (77.2, 77.8) | 3477 (70.5) | (69.2, 71.8) | 77518 (79.0) | (78.7, 79.2) |
|  | Outside major city | 1000 (31.2) | (29.6, 32.8) | 17261 (22.5) | (22.2, 22.8) | 1455 (29.5) | (28.2, 30.8) | 20655 (21.0) | (20.8, 21.3) |
| **SEIFA** | Lowest 50% | 1832 (57.2) | (55.5, 58.9) | 33595 (43.7) | (43.4, 44.1) | 2897 (58.7) | (57.4, 60.1) | 44419 (45.3) | (44.9, 45.6) |
|  | Highest 50% | 1372 (42.8) | (41.1, 44.5) | 43213 (56.3) | (55.9, 56.6) | 2035 (41.3) | (39.9, 42.6) | 53754 (54.8) | (54.4, 55.1) |

Figure 2 shows the proportion of young people with and without disability (living with parents at baseline) who were not co-residing with their parents five years later in the subsequent census. In 2021 (Figure 2, Panel A and Supplementary Table 1), a substantially lower proportion of people with disability (25.2% [95% CI 24.0, 26.4]) were no longer co-residing with parents compared to people without disability (49.1% [95% CI 48.8, 49.4]). An absolute difference of −23.9% (95% CI −25.2, −22.7) and relative inequality of 0.51 (95% CI 0.48,0.54) was observed between people with and without disability. The differences in no longer co-residing with parents between people with and without disability remained regardless of sex, age, engagement status, location, and individual income. The biggest inequalities between people with and without disability were observed for those who lived outside major cities (absolute −32.0% [95% CI −34.5, −29.6]; relative 0.48 [95% CI 0.44, 0.52]), those aged 25-29 years (absolute −39.3% (95% CI −42.5, −36.1); relative 0.41 [95% CI 0.36, 0.45]), and those who had individual income in the highest 60% (absolute −31.8% [95% CI −35.9, −27.7]; relative 0.53 [95% CI 0.47, 0.59]). The subgroup of people most likely to still be living with parents were males with disability, with 20.9% (95% CI 19.5, 22.3) no longer co-residing with parents and people with disability who were aged 15-19 years in the first wave (22.5% [96% CI 20.8, 24.3] no longer living with parents). A higher proportion of females with and without disability were no longer living with their parents (32.1% [95% CI 30.0, 34.2]) versus 53.2% [95% CI 52.7,53.6]). Similar patterns of transitions out of the parental home and inequalities between people with and without disability were seen in the 2011-2016 analysis, suggesting no evidence of improvements over time (Figure 2).

**Figure 2:**
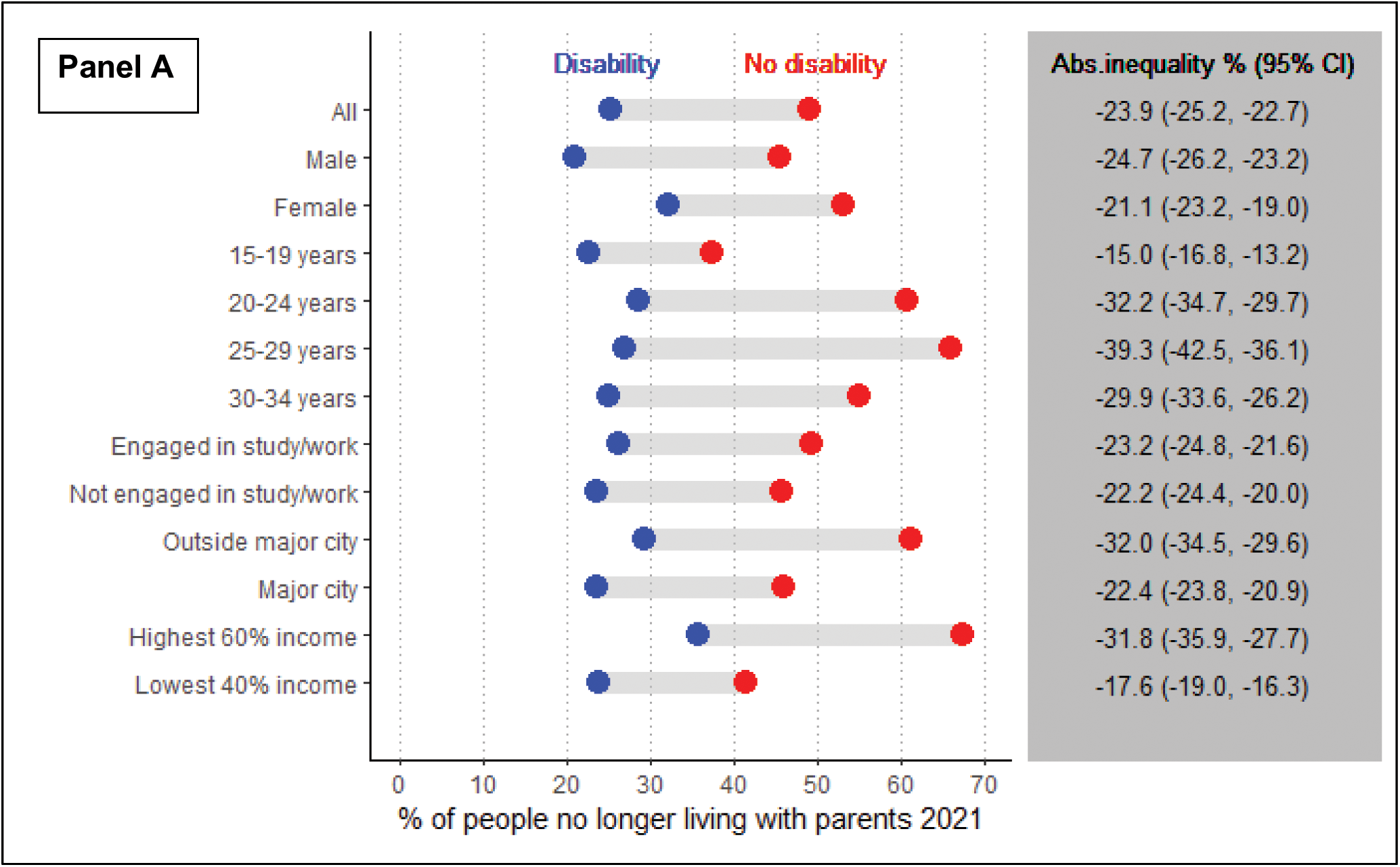
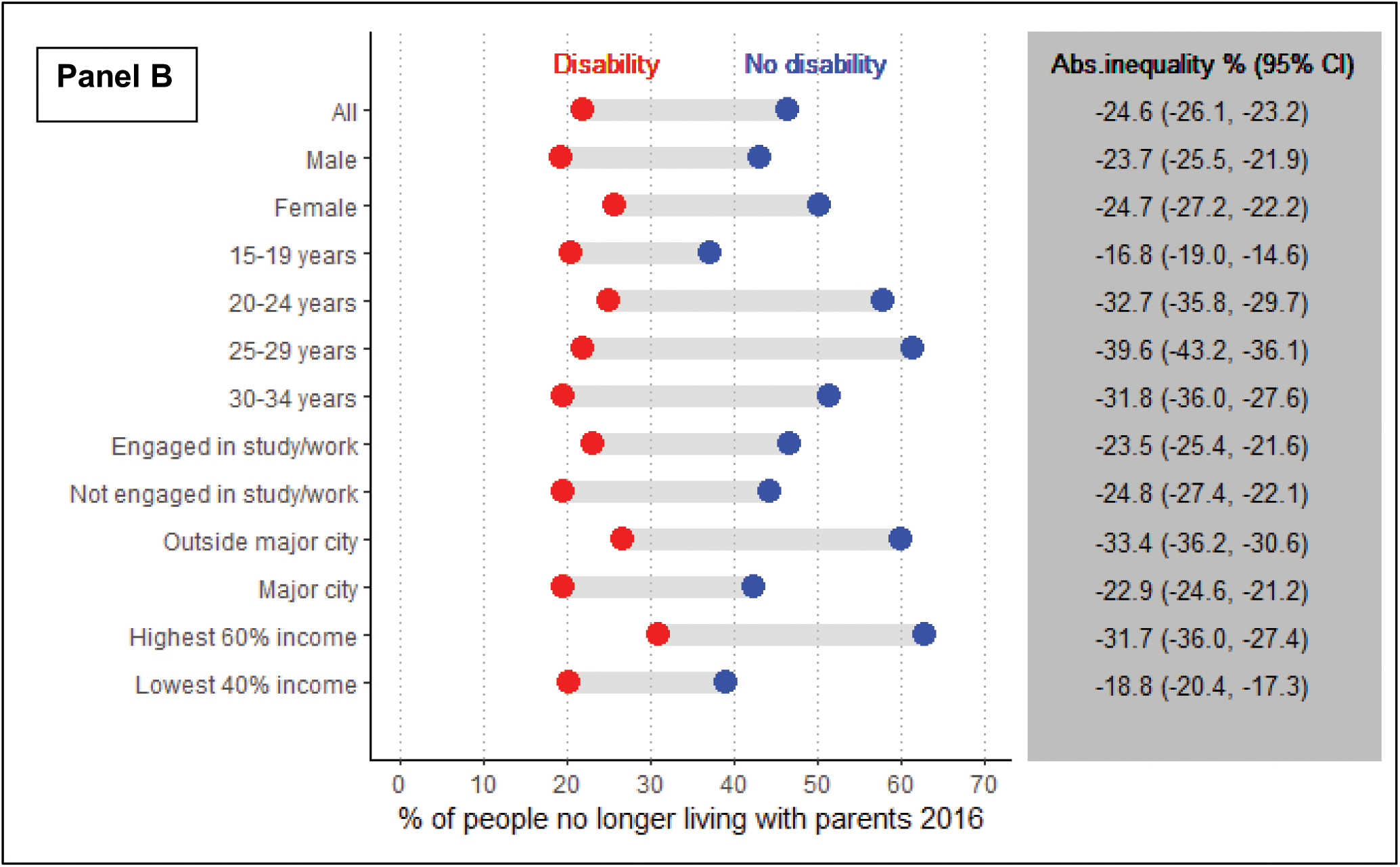
Proportion and absolute inequality of people with and without disability no longer living with parents in 2021 (Panel A) and 2016 (Panel B)

## Discussion

This study has found consistent and substantial inequalities in housing transitions between young people with and without disability. For people who were living with their parents at baseline, we found only a quarter of young people with disability moved out of the parental home compared to almost half of young people without disability. These inequalities held across all subgroups including sex, age, income, location and engagement in employment or education, but were greater for certain subgroups. Young people without disability who were aged 25-29 years were less than half as likely to have moved out of the parental home than people with disability in this age group. For those living outside of major cities, young people with disability were half as likely to have moved out of the parental home than their peers without disability. Finally, there were large inequalities among those with income in the highest 60%, with an absolute difference of 32% between young people with and without disability. The gaps remained consistent in the two sets of analyses, suggesting that there has not been an improvement over time.

To our knowledge, this is the first study to describe transitions out of the parental home for people with and without disabilities in Australia, and internationally. Our findings for people without disabilities align with a report from the Australian Institute of Family Studies (using 2016 census data) that found that 43% of 20-24 year olds, and 17% of 25-29 year olds were still living in the family home (33). For people with disabilities, the Australian Institute of Health and Welfare (AIHW) reported that 63% of people with disability aged 18-24 years were still living in their parental home, compared to 69% of people without disability (7). This contrasts to our findings, which reveal bigger differences between people with and without disabilities. This may be because we examined housing transitions using longitudinal rather than cross sectional data, allowing for within-person comparisons. Leaving the parental home is often considered a key milestone in the transition to adulthood, and an important indicator of independence. Remaining living in the parental home may indicate lack of choice and control about where young people live and with whom. We found that those people with disability who have higher incomes and are engaged in education or employment remain living with their parents at high rates, suggesting the barriers are more than financial. These could relate to supports required to live independently or lack of accessible housing options. The Royal Commission into Violence, Abuse, Neglect, and Exploitation of People with Disability (DRC) Report identified a range of barriers preventing people with disability in Australia from living in suitable housing and being able to choose where they live, with whom, and how. These barriers include tenancy insecurity, difficulties accessing social housing, difficulty finding accessible housing, and poor living conditions in supported boarding houses (34).

Our findings contribute to international literature on housing pathways by demonstrating inequalities in leaving the parental home, even within a formal disability income and support system. Importantly, this study shows there has been no improvement over time in transitions from the parental home for people with disability. This is despite introduction of a major reform in disability policy – the NDIS – between the two time periods. There are important implications arising for people with more severe or profound disability, who are eligible for NDIS home and living support. Young people with severe disability and complex support needs may remain living with their parents due to their need for support with tasks of daily living even though NDIS home and living supports are available. This suggests there may be issues with implementation of NDIS home and living support. Remaining in the parental home can be challenging as parents of adult children with severe disability age, with parents often providing support whilst also dealing with their own aging and reduced capabilities (32). One Australian study highlights that parents have to deal with their own ageing alongside providing a range of supports to adult offspring with intellectual disability. With limited safe and affordable options for their children, it is difficult to plan for the future (35). If not living with their parents, young people with severe or profound disability are often left with little choice but to live in segregated housing such as group homes or residential aged care. As highlighted by the DRC these settings are associated with increased risks of abuse and neglect and are at odds with Australia’s commitments as a signatory to the UNCRPD for people with disability to have “freedom from exploitation, violence and abuse” and the right to “living independently and being included in the community” (1).

Although home and living funding available on the NDIS is important for addressing accessibility and provision of supports, this does not address the broader housing affordability crisis. The majority (∼70%) of young people with disability do not receive NDIS funding so options are further limited. Australia, like other countries, is currently experiencing a cost-of-living crisis – driven mainly by inflation. This has resulted in increases in the costs of food, housing, utilities, and other basic necessities, and with no commensurate increase in wages. Low rental vacancy rates, and rental price increases make it difficult for many to find and afford suitable housing. Recent data illustrates how these pressures are intensifying. Reports from the AIHW have shown that the housing crisis has exacerbated challenges for people with disability, with already prolonged average wait times for public housing increasing by 134 days between 2020-21 and 2023-24, resulting in average wait times of 547 days (36). Compared to other OECD countries, Australia lags behind in public housing with this making up only 4.1% of all dwellings, compared to, for example, 16.7% in the United Kingdom (37–39). This means demand far outweighs supply and drives long waiting lists. Further, a recent longitudinal analysis of the HILDA Survey data found that the proportion of people living in unaffordable housing has increased over the last two decades – from 2.9% in 2003 to 4.6% in 2022 for people without disability and 7.3% in 2003 to 10.4% in 2022 for people with disability (2). Rising housing costs and declining affordability have particular implications for young people seeking to leave the family home. Shared rental housing is often a first step out of the family home, and some people are eligible (e.g. people in receipt of JobSeeker or Youth Allowance payments) to receive rent assistance through the Commonwealth Rent Assistance scheme. However, there is a large gap between actual rents and rent assistance which may prevent people from moving out. Policy debates often highlight the inadequacy of statutory incomes and associated housing assistance particularly for individuals who receive Youth Allowance, relative to housing costs (25).

These pressures have huge impacts on young people, with Orygen’s Youth Survey (of 15-19 year olds) finding 50% of those experiencing financial concerns or financial stress lived in unstable housing (40). More needs to be done to support young people, particularly those with disability, in terms of financial support and provision of housing options. It is also important to consider the location of housing provision to ensure suitable options are well-located for young people (easy access to amenities).(41) The inequalities identified in this paper underscore the need for tailored housing policies and supports for young people with disability to help with the transition to independent living. This aligns with broader current debates about housing choice for people with disability (27, 42).

There are also implications for people with disability’s mental health given that housing is a key social determinant of health (17). A large body of evidence shows that people with disability have poorer mental health than their peers without disability, in part driven by the social determinants of health and barriers to participation (43–45). Recent ABS data suggests people with disability experience psychological distress at nearly three times the rate of people without disability (46). It is possible that housing situations may influence the mental health of people with disability (3). However, there are few studies that have examined the mental health impacts of continuing to reside in the family home and living with parents. An Australian study found young adults who co-reside with parents had poorer mental health than those who lived independently.(8) Given a large proportion of young people with disability in our study were living with their parents, this raises concerns about potential poor mental health outcomes for this young cohort.

Although our study found no changes over time in young people living independently, the Covid-19 pandemic and lockdown measures may have influenced the findings in the analysis of 2021 Census data. Analyses from the Australian Institute of Family Studies found an increase in the numbers of young adults (20-21 years) returning to live with their parents at the start of the Covid-19 pandemic (47). However, in our study, there appeared to be very small increases in the proportion who moved out of the parental home for both people with or without disability between 2016 and 2021. Despite slight improvements, there is a need for policies to support people without disability as well in the context of the current cost of living crisis.

Although this study is based in Australia, the findings have relevance for other countries facing similar housing affordability pressures and policy shifts in disability supports. The persistence of inequalities over time suggests that without targeted housing and income interventions, disabled young people in many contexts may remain excluded from independent living.

### Implications for research

This paper has highlighted the inequalities in housing transitions for young people with disability. Further research is required to understand the reasons why young people with disability are more likely to remain living with their parents, including the structural and personal drivers of these inequalities, and the health, employment, and participation outcomes associated with different living arrangements. Future research should explore the housing needs and preferences of young people with disability to inform more responsive housing and support models. Investigating relationships and caregiving roles within households, such as whether parents are acting as unpaid carers, is also important.

Our study also considered disability in its broadest definition by including people with disability in either wave of the two sets of analyses. Further research could explore how transitions in disability status, e.g. through disability acquisition, may impact transitions out of the parental home for young people, as well as the impacts of long-term disability on this outcome.

### Strengths and limitations

This study’s main strength is the use of a large and unique dataset, the Australian Census Longitudinal Dataset (ACLD), to explore housing transitions for young people with and without disability. Given the size of the datasets (80,012 [2011], 103,105 [2016]), we were able to explore this issue in depth, which is not possible within other publicly available surveys nationally and internationally such as the HILDA Survey due to the small numbers of young people with disability.

The ACLD is a robust sample of the whole population of Australia, providing good representation and unlikely to be subject to selection bias. However, as with all Census data, there may be subpopulations that are missed due to living circumstances on Census night – such as those who are homeless (counts only) or those not residing in households. Because we restricted to people who were present in two consecutive ACLD waves, there are population dynamics that our sample may not capture – such as people who have migrated into Australia or out between the two time frames. Another strength of this paper was the engagement with young people with disability in the design of the study and interpretation of findings.

There are several limitations that need to be taken in to account when interpreting the results. Disability was measured using a set of four questions to derive ‘core activity need for assistance’ in self-care, communication or mobility due to a long-term health condition lasting 6 months or more. These questions underestimate the number of people with disability in the population with an estimated 5.8% in 2021 compared to 21.4% using SDAC in 2022 (28). The ABS states that the Census measure is comparable to the Survey of Disability Ageing and Carers concept of ‘profound or severe core activity limitation’ – i.e. mild or moderate limitations will not be picked up. Further, although four questions were used to generate the measure, it was provided as a binary variable within the ACLD based on the presence or absence of a core activity need for assistance. Thus we were unable to explore differences in housing transitions by severity of disability, level of support/assistance required, or by type. We were also unable to examine housing transitions for people with disability who do not have a core activity need for assistance, and thus cannot determine whether these individuals will have similar inequalities in housing transitions or be more similar to people without disability. Future research should use a broader disability flag to capture a more representative disability population (48).

The structure of the housing measure, based on household relationships, indicates that young people and their parents or grandparents live in the same household. Therefore, we cannot determine whether the young people are living with their parents/grandparents or vice versa. However, in this Australian context, the latter scenario would be relatively uncommon. We restricted our sample to people aged 15-34 years living with their parents or grandparents at baseline. This is a broad age group -people who are in the older end of this spectrum (e.g. over 30 years) who were still living with their parents at baseline are likely to still in this situation 5 years later. There is also a relatively large time period between censuses (5 years) and we do not have data in the intervening years to determine whether people may have left home and then returned. However, we wanted to examine long term transitions out of the parental home, so a large longitudinal dataset was required.

There was some missing data in our study. People with missing data were more likely to be Indigenous, or live in more disadvantaged areas, and have parents who were born outside of Australia. Approximately 10% of people aged 15-34 years living with their parents at baseline had missing data, which may introduce some bias and affect the representativeness of the findings. Finally, as with any data analysis using linked data, there is always the possibility of linkage errors or unlinked data. 24% of the ACLD sample in 2011 could not be linked to the 2016 Census and were thus excluded from the analysis (16% for the 2016-2021 ACLD). An analysis of linked vs unlinked data revealed of the 24% with unlinked data, there were some differences in characteristics (unlinked were more likely to be male, have a disability, identify as Indigenous, and live outside a major city), which may also introduce some bias and reduce how representative the findings are.

## Conclusions

This study provides important evidence on the stark inequalities in transitions out of the parental home between young people with and without disability in Australia. Young people with disability are half as likely to move out of the parental home compared to their peers without disability. These inequalities persist across key sociodemographic factors, including income, location, and engagement in work or study. The findings point to systemic gaps in housing policy, service provision, and support pathways for young people with disability. Addressing these barriers requires further research into the drivers of these inequalities, including the role of formal supports, household dynamics, and policy options. Ensuring equitable access to appropriate and preferred housing options is critical to enabling young people with disability to live independently.

## Data Availability

All data produced in the present work are held by the Australian Bureau of Statistics (ABS) in the secure DataLab platform. The authors do not have permission to share this data.

## Acknowledgements

This study uses the Person Level Integrated Data Asset (PLIDA), 2021, Australian Longitudinal Census Dataset, ABS DataLab. Findings based on use of PLIDA data.

This research was conducted on the lands of the Woi Wurrung language group of the Eastern Kulin nations, the traditional owners of the lands on Naarm, and the lands of the Yaegl people of the Yaygirr language group, the traditional owners of Yaegl Country. We pay our respects to Elders past, and present, and recognise Indigenous sovereignty was never ceded.

The researchers would also like to acknowledge Taranee Morney-Hype, Bella White, and Rosie Bogumil who are lived experience researchers on the Research Alliance for Youth Disability and Mental Health (RAY) programme and whose valuable insights shape our work.

## Appendices

### Supplementary tables

**Supplementary table 1:** Proportion of people who are no longer living with parents in 2021 (2016-2021 dataset)

|  |  | **Disability** |  | **No disability** |  | **Absolute inequality** | **Relative inequality** |
| --- | --- | --- | --- | --- | --- | --- | --- |
|  |  | N | % (95% CI) | N | % (95% CI) | % (95% CI) | RR (95% CI) |
|  | All | 1242 | 25.2 (24, 26.4) | 48209 | 49.1 (48.8, 49.4) | -23.9 (-25.2, -22.7) | 0.51 (0.48, 0.54) |
| **Sex** | Male | 632 | 20.9 (19.5, 22.3) | 23826 | 45.5 (45.1, 46.0) | -24.7 (-26.2, -23.2) | 0.46 (0.43, 0.49) |
|  | Female | 610 | 32.1 (30.0, 34.2) | 24383 | 53.2 (52.7, 53.6) | -21.1 (-23.3, -19.0) | 0.60 (0.56, 0.64) |
| **Age group** | 15-19 years | 497 | 22.5 (20.8, 24.3) | 19110 | 37.5 (37.1, 38.0) | -15.0 (-16.8, -13.2) | 0.60 (0.55, 0.65) |
|  | 20-24 years | 381 | 28.6 (26.3, 31.1) | 18425 | 60.8 (60.3, 61.3) | -32.2 (-34.7, -29.7) | 0.47 (0.43, 0.51) |
|  | 25-29 years | 211 | 26.8 (23.9, 30.1) | 7960 | 66.1 (65.3, 67.0) | -39.3 (-42.5, -36.1) | 0.41 (0.36, 0.45) |
|  | 30-34 years | 153 | 25.1 (21.8, 28.7) | 2714 | 55.0 (53.7, 56.4) | -29.9 (-33.6, -26.2) | 0.46 (0.39, 0.52) |
| **Engagement in study/work** | Engaged | 801 | 26.2 (24.7, 27.8) | 44574 | 49.4 (49.1, 49.7) | -23.2 (-24.8, -21.6) | 0.53 (0.50, 0.56) |
|  | Not engaged | 441 | 23.5 (21.7, 25.5) | 3635 | 45.7 (44.6, 46.8) | -22.2 (-24.4, -20.0) | 0.51 (0.47, 0.56) |
| **Location** | Outside major city | 426 | 29.3 (27.0, 31.7) | 12663 | 61.3 (60.6, 62.0) | -32.0 (-34.5, -29.6) | 0.48 (0.44, 0.52) |
|  | Major city | 816 | 23.5 (22.1, 24.9) | 35546 | 45.9 (45.5, 46.2) | -22.4 (-23.8, -20.9) | 0.51 (0.48, 0.54) |
| **Income** | Highest 60% | 190 | 35.7 (31.7, 39.8) | 19316 | 67.4 (66.9, 68.0) | -31.8 (-35.9, -27.7) | 0.53 (0.47, 0.59) |
|  | Lowest 40% | 1052 | 23.9 (22.7, 25.2) | 28893 | 41.6 (41.2, 41.9) | -17.6 (-19.0, -16.3) | 0.58 (0.55, 0.61) |

**Supplementary table 2:**
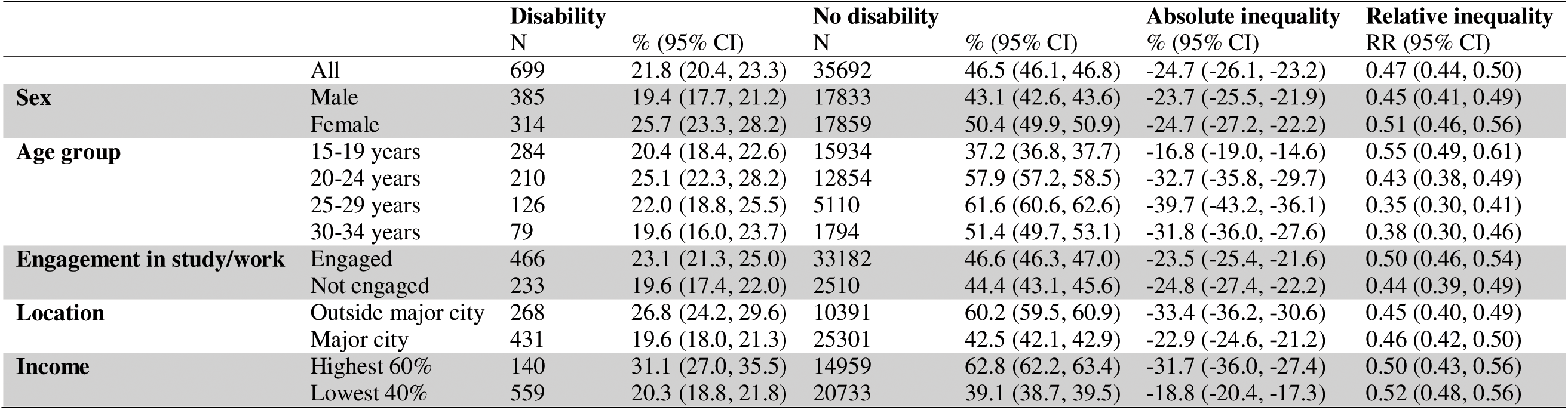
Proportion of people who are no longer living with parents in 2016 (2011-2016 dataset)

## References

1. United Nations Convention on the Rights of Persons with Disabilities. December 13,. 2006.

2. Sully A, Aitken Z, Bailie J, Bishop GM. Trends in disability-related inequalities in housing affordability in Australia, 2003 to 2022. Health & Place. 2025;95:103523.

3. Kavanagh AM, Aitken Z, Baker E, LaMontagne AD, Milner A, Bentley R. Housing tenure and affordability and mental health following disability acquisition in adulthood. Social Science & Medicine. 2016;151:225–32.

4. Aitken Z, Baker E, Badland H, Mason K, Bentley R, Beer A, Kavanagh AM. Precariously placed: housing affordability, quality and satisfaction of Australians with disabilities. Disability & Society. 2019;34(1):121–42.

5. Beer A, Faulkner DR. The housing careers of people with a disability and carers of people with a disability: Australian Housing and Urban Research Institute; 2008.

6. National Disability Data Asset. Identification of people with disability in linked administrative data for service use and outcomes reporting in housing. 2021.

7. Australian Institute of Health and Welfare. People with disability in Australia 2024 [Available from: https://www.aihw.gov.au/reports/disability/people-with-disability-in-australia/contents/people-with-disability.

8. Howard A, Li A, Bentley R. Parental co-residence and young adults’ mental health. PLOS ONE. 2023;18(11):e0294248.

9. Howden-Chapman P, Bennett J, Edwards R, Jacobs D, Nathan K, Ormandy D. Review of the Impact of Housing Quality on Inequalities in Health and Well-Being. Annu Rev Public Health. 2023;44:233–54.

10. Mason KE, Alexiou A, Li A, Taylor-Robinson D. The impact of housing insecurity on mental health, sleep and hypertension: Analysis of the UK Household Longitudinal Study and linked data, 2009–2019. Social Science & Medicine. 2024;351:116939.

11. Bess KD, Miller AL, Mehdipanah R. The effects of housing insecurity on children’s health: a scoping review. Health Promotion International. 2023;38(3):daac006.

12. Hock E, Blank L, Fairbrother H, Clowes M, Cuevas DC, Booth A, Goyder E. Exploring the impact of housing insecurity on the health and well-being of children and young people: a systematic review. 2023;11:13.

13. Bentley R, Baker E, Mason K, Subramanian SV, Kavanagh AM. Association Between Housing Affordability and Mental Health: A Longitudinal Analysis of a Nationally Representative Household Survey in Australia. American Journal of Epidemiology. 2011;174(7):753–60.

14. Baker E, Lester L, Mason K, Bentley R. Mental health and prolonged exposure to unaffordable housing: a longitudinal analysis. Social Psychiatry and Psychiatric Epidemiology. 2020;55(6):715–21.

15. Dotsikas K, Osborn D, Walters K, Dykxhoorn J. Trajectories of housing affordability and mental health problems: a population-based cohort study. Social Psychiatry and Psychiatric Epidemiology. 2023;58(5):769–78.

16. Singh A, Daniel L, Baker E, Bentley R. Housing Disadvantage and Poor Mental Health: A Systematic Review. American Journal of Preventive Medicine. 2019;57(2):262–72.

17. World Health Organization. WHO Housing and health guidelines 2018 [Available from: https://www.who.int/publications/i/item/9789241550376.

18. World Health Organization. International Classification of Functioning, Disability and Health (ICF) 2025 [Available from: https://www.who.int/standards/classifications/international-classification-of-functioning-disability-and-health.

19. Hochstenbach C, Howard A, Arundel R. Increasing Social and Spatial Inequalities in Parental Co-Residence. Tijdschrift voor Economische en Sociale Geografie. 2025;116(2):212–31.

20. Marinos S. More Australian adult children are living with their parents longer: Pursuit 2024 [Available from: https://pursuit.unimelb.edu.au/articles/more-australian-adult-children-are-living-with-their-parents-longer#:~:text=Having%20fun%20while%20they%20can,conditions%20aren’t%20allowing%20that.&text=%E2%80%9CPolicy%20action%20to%20make%20housing,who%20were%20.

21. Wilkins R, Vera-Toscano, E., Botha, F., Wooden, M., Trinh, T.,. The Household, Income and Labour Dynamics in Australia Survey: Selected Findings from Waves 1 to 20.: Melbourne Institute: Applied Economic & Social Research, the University of Melbourne; 2022.

22. Murinkó L. Pathways, background and outcomes of the transition to adulthood in the early 2000s in Hungary. Demográfia. 2018;61(5):59–103.

23. Wilkins R, Vera-Toscano, E., Botha, F., Dahmann, S.C,. The Household, Income and Labour Dynamics in Australia Survey: Selected Findings from Waves 1 to 19.: Melbourne Institute: Applied Economic & Social Research, the University of Melbourne; 2021.

24. Wiesel I, Laragy, C., Gendera, S., Fisher, K., Jenkinson, S., Hill, T., Finch, K., Shaw, W., Bridge, C.,. Moving to my home: housing aspirations, transitions and outcomes of people with disability 2015 [Available from: https://www.ahuri.edu.au/research/final-reports/246?utm_source=openai.

25. Parkinson S, Rowley, S., Stone, W., James, A., Spinney, A., Reynolds, M.,. Young Australians and the housing aspirations gap 2019 [Available from: https://www.ahuri.edu.au/research/final-reports/318?utm_source=openai.

26. Stone W, Rowley S, Parkinson S, James A, Spinney A. The housing aspirations of Australians across the life-course: closing the ‘housing aspirations gap’. AHURI Final Report. 2020(337).

27. Bennett S, Orban, H. Better safer more sustainable: How to reform NDIS housing and support. Grattan Institute Report. 2024.

28. Australian Bureau of Statistics. Disability, Ageing and Carers, Australia: Summary of Findings 2022 [Available from: https://www.abs.gov.au/statistics/health/disability/disability-ageing-and-carers-australia-summary-findings/latest-release.

29. Australian Bureau of Statistics. Characteristics of National Disability Insurance Scheme (NDIS) participants, 2019: Analysis of linked data 2021 [Available from: https://www.abs.gov.au/articles/characteristics-national-disability-insurance-scheme-ndis-participants-2019-analysis-linked-data.

30. Bishop GM, Kavanagh, A.M., Olney, S., Disney, G., Aitken, Z. . A comparison of the characteristics of people with disability in Australia according to whether they received National Disability Insurance Scheme (NDIS) funding. . Melbourne: The University of Melbourne; 2025.

31. Australian Bureau of Statistics. 2080.5 - Information Paper: Australian Census Longitudinal Dataset, Methodology and Quality Assessment, 2006-2016 2019 [Available from: https://www.abs.gov.au/ausstats/abs@.nsf/mf/2080.5.

32. Australian Bureau of Statistics. Socio-Economic Indexes for Areas (SEIFA), Australia methodology 2021 [Available from: https://www.abs.gov.au/methodologies/socio-economic-indexes-areas-seifa-australia-methodology/2021.

33. Australian Institute of Family Studies. Young people living with their parents 2019 [Available from: https://aifs.gov.au/research/facts-and-figures/young-people-living-their-parents.

34. Wiesel I. Mainstream housing and the Royal Commission into Violence, Abuse, Neglect and Exploitation of People with Disability. Research and Practice in Intellectual and Developmental Disabilities. 2024;11(1):29–38.

35. Walker R, Hutchinson C. Care-giving dynamics and futures planning among ageing parents of adult offspring with intellectual disability. Ageing and Society. 2019;39(7):1512–27.

36. Australian institute of Health and Welfare. Australian Disability Strategy 2021-2031: Outcomes Framework 2025 [Available from: https://www.aihw.gov.au/australias-disability-strategy/outcomes/inclusive-homes-and-communities/average-social-housing-wait-timehttps://www.aihw.gov.au/australias-disability-strategy/outcomes/inclusive-homes-and-communities/average-social-housing-wait-time.

37. OECD. OECD Affordable Housing Database - indicator PH4.2. Social rental housing stock 2024 [Available from: https://oe.cd/ahd.

38. Australian Institute of Health and Welfare. Housing assistance in Australia 2025 [Available from: https://www.aihw.gov.au/reports/housing-assistance/housing-assistance-in-australia/contents/summary.

39. Office for National Statistics. Subnational estimates of dwellings and households by tenure, England: 2023 2023 [Available from: https://www.ons.gov.uk/peoplepopulationandcommunity/housing/articles/researchoutputssubnationaldwellingstockbytenureestimatesengland2012to2015/2023.

40. Filia K, Teo, S.M., Gan, D., Browne, V., Baker, D., Menssink, J., Boon, B., Brennan, N., Di Nicola, K., Gao, C.X.,. Counting the cost of living – the impact of financial stress on young people. Insights from the 2023 Mission Australia Youth Survey. Orygen: Melbourne VIC and Mission Australia: Sydney, NSW.; 2024.

41. Alderton A, Aitken Z, Hewitt B, Dearn E, Badland H. Characteristics of geographic environments that support the health and wellbeing of young people with disability: A scoping review. Social Science & Medicine. 2025;370:117842.

42. Burke A, Walker, J.. Stoeckel, L., & Carey, F.,. Moving Out, Moving On: Beyond group homes for NDIS participants. . Summer Foundation; 2025.

43. Cree RA. Frequent mental distress among adults, by disability status, disability type, and selected characteristics—United States, 2018. MMWR Morbidity and mortality weekly report. 2020;69.

44. Stickley A, Kondo N, Roberts B, Kizilova K, Waldman K, Oh H, et al. Disability and psychological distress in nine countries of the former Soviet Union. Journal of affective disorders. 2021;292:782–7.

45. Aitken Z, Bishop GM, Disney G, Emerson E, Kavanagh AM. Disability-related inequalities in health and well-being are mediated by barriers to participation faced by people with disability. A causal mediation analysis. Soc Sci Med. 2022;315:115500.

46. AIHW. Mental health services in brief 2019. 2019.

47. Australian Institute of Family Studies. Young adults returning to live with parents during COVID-19. 2023.

48. Aitken Z, Walmsley S, G MB, Badji S, Fortune N. Methods used to construct disability indicators in linked administrative datasets: a systematic scoping review. Popul Health Metr. 2025;23(1):22.

